# National Changes in Diabetes Care Practices during the COVID-19 Pandemic: Prospective Study of US Adults

**DOI:** 10.1101/2023.08.06.23293722

**Authors:** Kushagra Vashist, Saria Hassan, Mary Beth Weber, Rakale C. Quarells, Shivani A. Patel

## Abstract

**Background:** There is a lack of nationally representative prospective data on the impact of the COVID-19 pandemic on diabetes care and management in adults with type 2 diabetes. We examined changes in diabetes care and management practices before and after the onset of the COVID-19 pandemic.

**Methods:** Using the National Health Interview Survey, we analyzed data from 870 adults living with type 2 diabetes who were interviewed in 2019 and re-interviewed between August and December 2020. Exposure to the COVID-19 pandemic was defined by year of survey (2019, pre-pandemic; 2020, pandemic). We estimated percent change in past year blood sugar check by a health professional and current use of blood sugar lowering medication overall and by sociodemographic subgroups.

**Results:** Receiving an annual blood sugar test fell by -3.3 percentage points (pp) (95% CI -5.7, -1.0), from 98.3% in 2019 to 95.0% in late 2020. The reduction in annual blood glucose testing was largely consistent across socio-demographic groups and was particularly pronounced among adults not working and adults aged 65 years and older. In the same time period, current use of diabetes medications increased by +3.8 pp (0.7, 6.9), from 85.9% to 89.7%. The increase in medication use was most pronounced among individuals aged 40-64-year old, employed, and those living in large central metropolitan areas.

**Conclusions:** Nationally, adults with Type 2 diabetes reported a reduction in annual blood glucose testing by a health professional and an increase in diabetes medication usage during the COVID-19 pandemic. If sustained after the end of the COVID-19 public health emergency, these changes have implications for national diabetes management and care.

## Introduction

Diabetes is a complex chronic condition requiring routine monitoring and regular medication. At minimum, the American Diabetes Association (ADA) recommends having an annual blood sugar check ^1^ and when needed, pharmacologic treatments such as metformin or other agents, including combination therapy ^2^ in adults with diabetes. There has been much concern that the COVID-19 pandemic has impacted diabetes care. Disruptions to health services ^3, 4^, changes to routine care ^4^, and fear of getting severely ill from the virus ^5^ may have prevented some individuals living with diabetes from getting the care they need. Lack of access to health services has been associated with diabetes-related complications such as cardiovascular disease, kidney disease, neuropathy, or blindness ^5^. Conversely, access to healthcare has been associated with controlled levels of blood glucose, blood pressure, and blood lipids ^6^, which collectively prevent the onset of diabetes-related complications ^7, 8^.

Several studies indicate that that people with diabetes experienced challenges with managing their disease during the pandemic. For example, an online community-based survey indicated that 25% of people needing an insulin pump or continuous glucose monitoring supplies had delays or difficulties in obtaining them and that 1 in 6 persons with diabetes had difficulty needing insulin, as of April 2020 ^9^. However, these data do not provide a clear picture regarding how individuals’ diabetes care and management have changed in response to the pandemic.

Identifying the indirect impact of the pandemic is critical to understanding the needs and requirements of those living with diabetes for future public health emergencies and the state of diabetes care as we move into post-public health emergency recovery. Using prospective data from a nationally representative sample of adults interviewed in 2019 and re-interviewed in 2020, we examined changes in annual blood sugar checks and use of current medication to lower blood sugar — two key diabetes care and management practices— before and after the onset of the COVID-19 pandemic among people living with type 2 diabetes.

## Materials and METHODS

### Data and sample

The National Health Interview Survey (NHIS) is a representative household interview survey which collects information on the health of non-institutionalized US population across the 50 states through a complex multistage design. To understand the ramifications of the COVID-19 pandemic, NHIS re-interviewed a sample of adults in August-December 2020 from 2019.

This longitudinal sample started with 21,161 adults from non-Medical Expenditure Panel Survey that completed or partially completed a 2019 Sample Adult Interview ^10^. After satisfying the eligibility criteria, this number was reduced to 20,827. Of these eligible sample adults, 10,415 adults interviewed in August-December 2020. We merged the publicly available 2019 NHIS adults sample data with their follow up data in 2020 to conduct analysis of change in diabetes outcomes ^10^. The analysis was restricted to adults with self-reported type 2 diabetes. Of the 973 longitudinal sample respondents living with type 2 diabetes aged 18-96 years, 10.6% were excluded from the analysis because of missing information on a study outcome or covariate, resulting in an unweighted analytic sample size of 870.

NHIS is approved by the Research Ethics Review Board of the National Center for Health Statistics and the U.S Office of Management and Budget. All NHIS respondents provided oral consent prior to participation ^11^. This study used publicly available data containing no personal identifying information and is therefore deemed not human subjects research by the Emory IRB.

### Variables and description

Our two outcomes included having blood sugar checked by a doctor, nurse, or other health professional within the past 12 months and taking medication to lower blood sugar for diabetes control (either oral/hypoglycemic agents or insulin).

#### Socio-demographic characteristics

Demographics included sex (male/female), age (divided into three groups; 18-39, 40-64, and 65 and older), race/ethnicity (as non-Hispanic White, non-Hispanic Black, Hispanic, and Other), smoking status (former/never smoker and current everyday/someday smoker) and marital status (married or living with a partner). Socioeconomic characteristics included educational attainment (classified as having less than bachelor’s degree or bachelor’s degree or more), urbanicity (categorized as large central/fringe metropolitan [population > 1 million], medium and small metropolitan, and nonmetropolitan), employment status (full-time and/or part-time employed, retired, unemployed/not working), and insurance status (no insurance, private insurance, Medicare insurance, Medicaid insurance, and other public insurance). Types of insurance were mutually exclusive and individuals with multiple health insurance were assigned a single insurance in the following order: those with private insurance included individuals who only had private insurance or a combination with any other coverage; next, those with Medicare insurance included having Medicare alone or a combination with any other public insurance; and lastly, those having Medicaid insurance included persons who had Medicaid insurance alone or in combination with any non-Medicare, public insurance.

### Statistical analysis

We first described the distribution of socio-demographic characteristics of respondents at baseline in 2019.

We estimated the prevalence of annual blood sugar checked and taking medication to lower blood sugar for diabetes control in 2019. Next, we estimated the average within-person change in each outcome from 2019 to 2020. Changes were estimated for the total sample as well as by age, sex, race, smoking status, marital status, urbanicity, insurance type, education level, and employment status. We computed the unadjusted change in each stratum, as well as the marginally adjusted change accounting for other sociodemographic groups in the analysis.

All data were analyzed using SAS version 9.4 and SUDAAN version 11.0.1, accounting for the complex survey design.

## RESULTS

The distribution of the sociodemographic characteristics of adults with type 2 diabetes at baseline (2019) are shown in Table 1.

**Table 1.** Distribution of socio-demographic characteristics among those living with type 2 diabetes, baseline interview, January 2019-December 2019.

|  | Unweighted n | Weighted % (95% CI) |
| --- | --- | --- |
| **Characteristics** |  |  |
| Overall | 870 | 100.0 (-) |
| **Sex** |  |  |
| Male | 405 | 49.9 (45.4, 54.4) |
| Female | 465 | 50.1 (45.6, 54.6) |
| **Age** |  |  |
| 18-39 | 20 | * |
| 40-64 | 365 | 50.7 (46.2, 55.3) |
| 65 and older | 485 | 45.1 (40.2, 50.0) |
| **Race/ethnicity** |  |  |
| Non-Hispanic White | 596 | 59.0 (53.9, 63.9) |
| Non-Hispanic Black | 111 | 13.2 (10.2, 16.9) |
| Hispanic | 95 | 16.7 (12.6, 21.7) |
| Other | 68 | 11.1 (8.1, 15.0) |
| **Smoking status** |  |  |
| Former/never smoker | 772 | 89.6 (86.7, 92.0) |
| Current every day/some day smoker | 98 | 10.4 (8.0, 13.3) |
| **Education** |  |  |
| Less than 4 years of college | 633 | 81.0 (77.8, 83.9) |
| 4 years of college or more | 237 | 19.0 (16.1, 22.2) |
| **Employment status** |  |  |
| Employed | 307 | 39.9 (35.6, 44.5) |
| Retired | 403 | 39.0 (34.5, 43.7) |
| Not working† | 160 | 21.0 (17.5, 25.1) |
| **Marital status** |  |  |
| Married or living with a partner | 423 | 63.0 (58.8, 67.1) |
| Unmarried | 447 | 37.0 (32.9, 41.2) |
| **Insurance type‡** |  |  |
| Not insured | 32 | 6.6 (4.3, 10.0) |
| Private insurance | 480 | 53.2 (48.5, 58.0) |
| Medicare | 281 | 28.2 (24.2, 32.6) |
| Medicaid | 62 | 10.0 (6.6, 14.8) |
| Other | 15 | * |
| **Urbanicity** |  |  |
| Large central/fringe metropolitan | 410 | 49.7 (43.8, 55.5) |
| Medium and small metropolitan | 278 | 29.9 (24.9, 35.5) |
| Nonmetropolitan | 182 | 20.4 (16.1, 25.5) |

\*Percentages suppressed for n<30
†Consists of persons unemployed, unable to work for health reasons/disabled, taking care of house or family, going to school, and not working because of other reasons
\*Types of insurance coverage are mutually exclusive. Persons with multiple types of health insurance were assigned to the first appropriate category in the following order: privately-insured, Medicare-insured, and Medicaid-insured

Table 2 shows the prevalence of diabetes care practices in 2019 and the unadjusted and adjusted average percent change from 2019 to 2020 Nationally, the percentage receiving an annual blood sugar check fell by an average of 3.3 percentage points (pp) (95% Confidence Interval [CI] -5.7, -1.0) from 98.3% in 2019 to 95.0% in Aug-Dec 2020. The reduction in annual blood sugar checking was largely consistent across sociodemographic groups, and was particularly pronounced among males (-4.7 pp [CI: -8.4, -1.1]), those not working (-8.1 pp [CI:-14.4, -1.8]) and 65 years and older (-4.9 pp [CI: -9.2, -0.7]). The percentage of participants taking medication to lower blood sugar increased by +3.8 pp (CI: 0.7, 6.9), from 85.9% to 89.7% from 2019 to 2020. The increase in anti-glycemic medication was most pronounced among those living in large central metropolitan area (5.8 pp [CI: 0.6, 11.0]) and 40-64 years old (4.8 pp [CI: 0.4, 9.2].

**Table 2.**
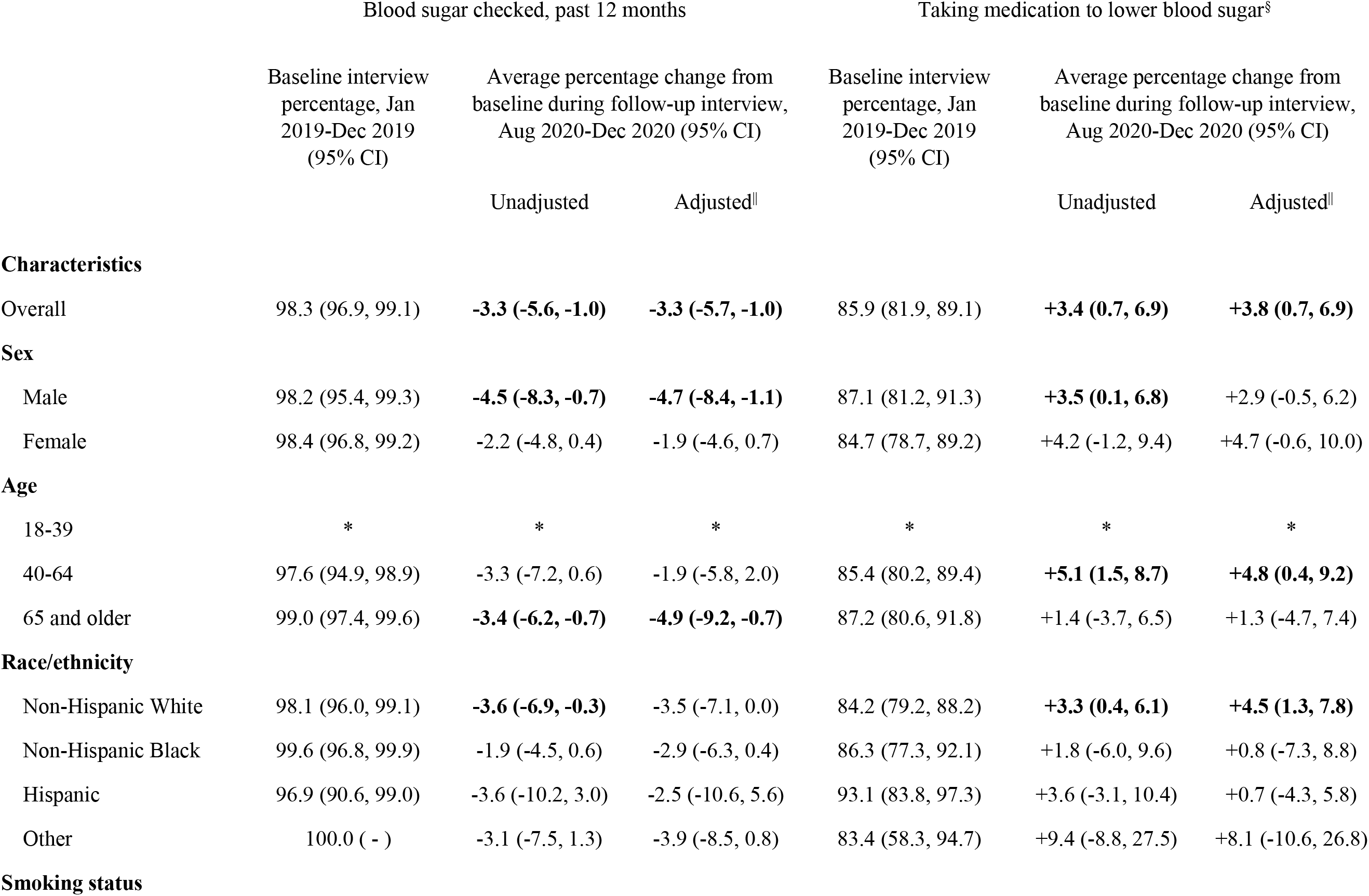
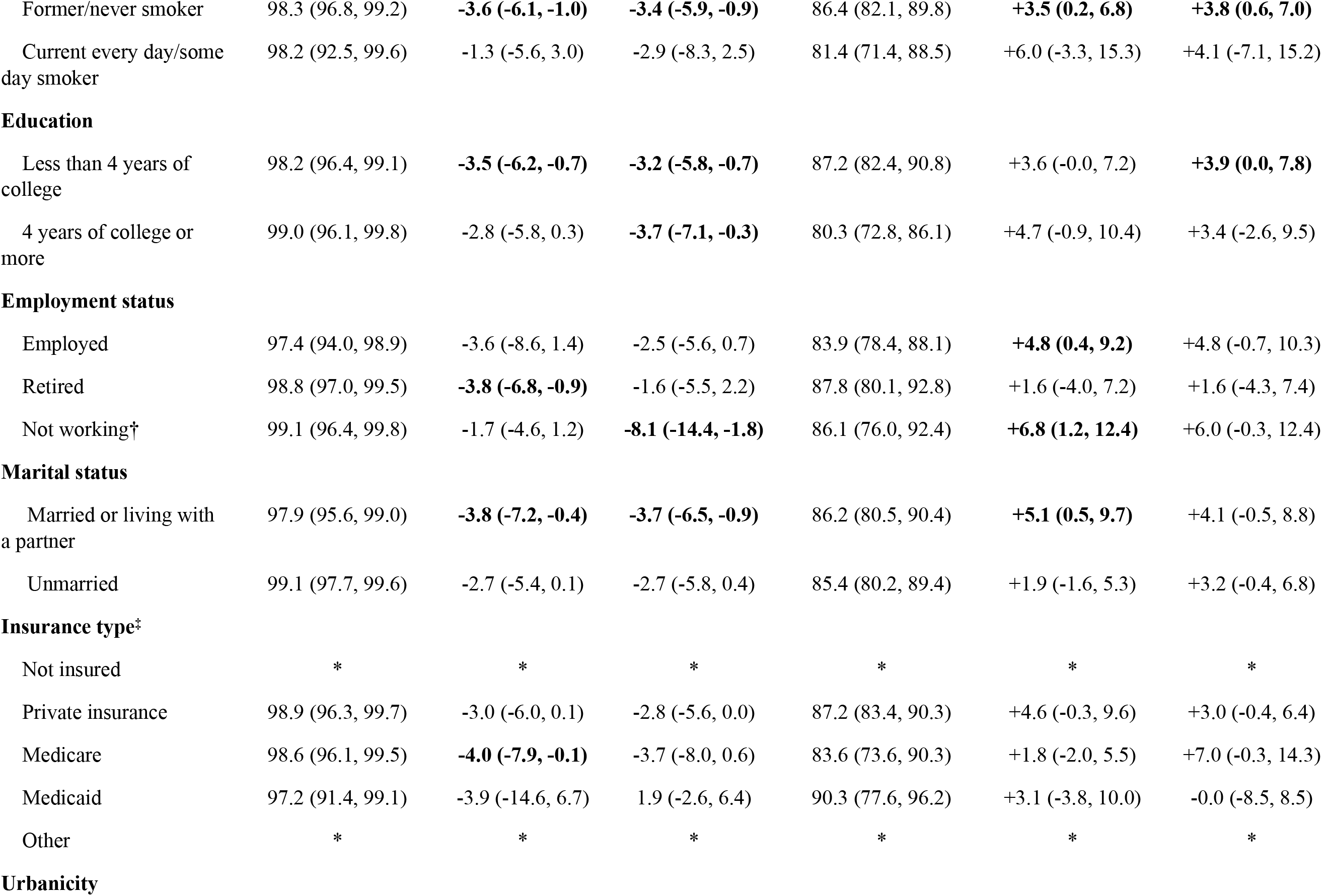
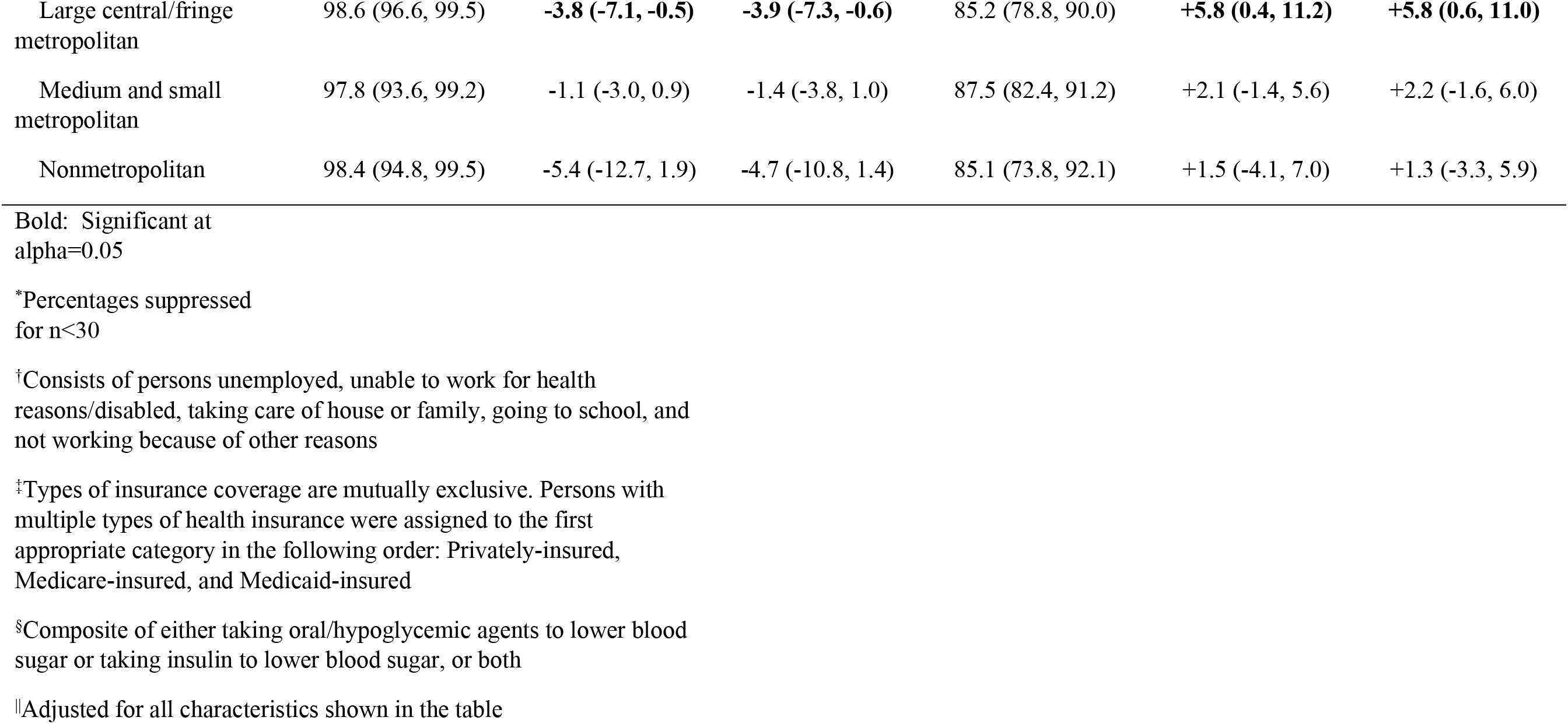
Taking medication to lower blood sugar and having blood sugar checked, past 12 months at baseline and average percentage change at follow-up among those living with type 2 diabetes.

## DISCUSSION

To our knowledge, this is the first study which uses nationally representative, community-based prospective data to examine within-person changes in diabetes care practices associated with the COVID-19 pandemic. Among adults with type 2 diabetes, having blood sugar checked by a health professional decreased during the pandemic. On the other hand, taking medications to lower blood sugar for diabetes control increased. The findings suggest that the pandemic had a negative impact on diabetes care but an unclear impact on diabetes medication. The increase in diabetes medication usage may suggest a positive impact on diabetes control. On the other hand, it may have been that blood sugar levels worsened during the pandemic, leading more individuals to take medication for diabetes control.

Reasons for decreased testing during the pandemic may have been due to disruption of health services ^3, 4^, changes in routine care ^4^, and fear of getting ill from the COVID-19 infection when going to the doctor ^5^. Although reductions in testing were consistent across socio-demographic subgroups, changes differed in magnitude for some. We saw a greater decrease in blood sugar checked in individuals 65 years and older compared to other age groups. Older individuals are at a higher risk of getting severely ill from COVID-19 ^12^, which may have been a factor in deterring them from visiting a health professional for testing or health professionals might have encouraged telehealth only visits for this high risk group. Adults who were not working at the 2019 baseline reported the largest decrease in having blood sugar checked (-8.1%), compared to other employment groups. The not working employment group largely consisted of persons unemployed, unable to work for health reasons/disabled, and taking care of house or family, and it may have been that these individuals were restricted in their ability to seek healthcare for the very reasons they were classified as not working.

Respondents reported an average increase in use of medications to lower blood sugar in August-December 2020 compared to 2019. This finding is consistent with reports from a previous study using an online survey, which showed that among respondents with type 2 diabetes who reported a change in the intake of diabetes medication, a greater proportion of individuals took medications more regularly during the pandemic compared to before the pandemic ^13^. The increase in diabetes usage during the early pandemic could be explained by one of two scenarios: 1. Patients were previously prescribed a diabetes lowering medication but had not started it or not taking it for some reason; or 2. Patients previously not eligible for diabetes medication based on their blood sugar values, were now eligible for blood sugar lowering medication. Alternatively, persons may have been more diligently checking their blood sugar at home, identified rising blood sugars, contacted their provider, and received newly prescribed blood sugar lowering medication. It is noteworthy that individuals with diabetes were able to get refills on prescriptions without visiting the doctor in 2020, whereas they had to return to their doctor to get refills by 2021.

In our analysis, we found that those living in large central metropolitan areas and in the 40-64 years old age group reported a higher increase in the intake of medication to lower blood sugar in 2020 compared to those living in less urban areas or younger/older age groups.

Individuals living in urban areas may have better infrastructure to obtain medications ^14^, better access to telehealth services ^15^, and be able to better afford medications compared to rural places^14^. Individuals aged 40-64 reported an increase in medication to lower blood sugar. These individuals may also be more likely to have access to internet and telehealth services ^16^ to enable discussion with providers on the need for medication despite not being able to visit the provider.

Broadly, our findings regarding both medication and testing are consistent with other national studies examining diabetes testing and medication usage during the pandemic. A previous study using electronic medical records in the United States during the same time period found a decrease outpatient visits and HbA1c testing during the pandemic, without evidence of reduced medication fills or glucose control ^17^. Considering serial cross-sectional data from NHIS respondents, the prevalence of blood glucose checking in US adults with diabetes was 96.8% in 2019 compared with 94.2% in 2021, suggesting that the drop in glucose checking by health professionals was sustained even in the second year of the pandemic ^18^. Similarly, reductions in diabetes testing have also been reported in England ^19^. Reductions in HbA_1C_ testing is of concern due the importance of glucose monitoring for clinical treatment decisions and feedback to patients on diabetes management ^19^.

Our study has several strengths. First, we used nationally representative prospective data to quantify diabetes care practices among the same individuals with type 2 diabetes before (2019) and after the onset of the pandemic (2020). Second, we examined changes in taking medication for diabetes control and having annual blood sugar checked from 2019 to 2020 by key sociodemographic groups, highlighting significant changes. Several limitations of this study should be acknowledged. There may have been overlapping time periods between 2019 and 2020 for responses to having blood sugar checked by a health professional in the past 12 months. We were not able to assess whether changes in diabetes testing and medication resulted from changes in diabetes control due to lack of laboratory data.

## CONCLUSIONS

Using data from prospectively followed adults drawn from a nationally representative sample, we found a reduction in annual blood sugar checks by healthcare providers and an increase in reporting of taking medication to lower blood sugar in 2020 compared to 2019, among adults with type 2 diabetes. Despite a reduction in testing, there was an increase in those taking medication for diabetes. The increase in medication warrants further examination. Whether and to what extent these changes were sustained after the first year and will set a “new normal” for diabetes care and management is not fully clear. Understanding the indirect impact of the pandemic on diabetes care is critical to equitably addressing the needs of those living with diabetes in future public health emergencies and their aftermath. The need to intervene on communities and individuals at greater risk is necessary to reduce the impact of public health emergencies and ensure equitable access to recommended diabetes care and management practices.

## Data Availability

All data are held in public repository at CDC's website on NHIS data. https://www.cdc.gov/nchs/nhis/2020nhis.htm https://www.cdc.gov/nchs/nhis/2019nhis.htm

https://www.cdc.gov/nchs/nhis/2019nhis.htm

## Funding and assistance

KV, MBW, RCQ, and SAP were supported by grant number P30DK111024-05S2 from the National Institute of Health under the RADx® Underserved Populations (RADx-UP) Initiative, and MBW is supported by P30DK111024. SH is supported by a grant from the National Institute of Health/National Heart Lung and Blood Institute (K23HL152368).

## Conflict of interest

The authors declare that there are no relevant conflicts of interest.

## Author contributions

KV and SAP conceptualized the study. KV conducted the statistical analysis and wrote the first draft of the manuscript. SAP provided supervision of the study and input into study design. All authors contributed to the interpretation of data. SH, MBW, RCQ, SAP reviewed and provided substantive revisions to the manuscript. All authors approved the final version of the manuscript. KV is the guarantor of this work and, as such, had full access to all the data in the study and takes responsibility for the integrity of the data and the accuracy of the data analysis.

## Notes

### Competing Interest Statement

The authors have declared no competing interest.

### Author Declarations

This study used publicly available data containing no personal identifying information and is therefore deemed not human subjects research by the Emory IRB.

